# A short therapeutic regimen based on hydroxychloroquine plus azithromycin for the treatment of COVID-19 in patients with moderate disease. A strategy associated with a reduction in hospital admissions and complications

**DOI:** 10.1101/2020.06.10.20101105

**Authors:** José A. Oteo, Pedro Marco, Luis Ponce de León, Alejandra Roncero, Teófilo Lobera, Valentín Lisa

## Abstract

The new SARS-CoV-2 infection named COVID-19 has severely hit our Health System. At the time of writing this paper no medical therapy is officially recommended or has shown results in improving the outcomes in COVID-19 patients. With the aim of diminishing the impact in Hospital admissions and reducing the number of medical complications, we implemented a strategy based on a Hospital Home-Care Unit (HHCU) using an easy-to-use treatment based on an oral administration regimen outside the hospital with hydroxychloroquine (HCQ) plus azithromycin (AZM) for a short period of 5 days.

**Patients and methods:** Patients ≥ 18 years old visiting the emergency room at the Hospital Universitario San Pedro de Logroño (La Rioja) between March, 31^st^ and April, 12^th^ diagnosed with COVID-19 with confirmed SARS-CoV-2 infection by a specific PCR, as follows: Patients with pneumonia (CURB ≤ 1) who did not present severe comorbidities and had no processes that contraindicated this therapeutic regime. Olygosimptomatic patients without pneumonia aged ≥ 55 years. Patients ≥ 18 years old without pneumonia with significant comorbidities. We excluded patients with known allergies to some of the antimicrobials used and patients treated with other drugs that increase the QTc or with QTc >450msc. The therapeutic regime was: HCQ 400 mg every twice in a loading dose followed by 200 mg twice for 5 days, plus AZM 500 mg on the first day followed by 250 mg daily for 5 days. A daily telephone follow-up was carried out from the hospital by the same physician.

The end-points of our study were: 1.- To measure the need for hospital admission within 15 days after the start of treatment. 2.- To measure the need to be admitted to the intensive care unit (ICU) within 15 days after the start of the treatment. 3.- To describe the severity of the clinical complications developed. 4.- To measure the mortality within 30 days after starting treatment (differentiating if the cause is COVID-19 or something else). 5.-To describe the safety and adverse effects of the therapeutic regime.

**Results:** During the 13 days studied a total of 502 patients were attended in the emergency room due to COVID-19. Forty-two were sent at home; 80 were attended by the HHCU (patients on this study) and 380 were admitted to the Hospital. In our series there were a group of 69 (85.18%) patients diagnosed with pneumonia (37 males and 32 females). Most of them, 57 (82.60%) had a CURB65 score of <1 (average age 49) and 12 (17.40%) a CURB score of 1 (average age 63). Eighteen (22.50%) of the pneumonia patients also had some morbidity as a risk factor. 11 patients (13.75%) without pneumonia were admitted to the HHCU because comorbidities or age ≥ 55 years. Six patients with pneumonia had to be hospitalized during the observation period, 3 of them because side effects and 3 because of worsening. One of these patients, with morbid obesity and asthma, had clinical worsening needing mechanical ventilation at ICU and developed acute distress respiratory syndrome. With the exception of the patient admitted to the ICU, the rest of the patients were discharged at home in the following 8 days (3 to 8 days).

Twelve patients (15%), 11 of whom had pneumonia, experienced side effects affecting mainly the digestive. In another patient a QTc interval prolongation (452 msc) was observed. In total 3 of these patients had to be admitted in the Hospital, 2 because of vomiting and 1 because a QTc interval lengthening. None of the patients needed to stop the HCQ or AZM and all the 80 patients finished the therapeutic strategy. From the group without pneumonia only a patient developed diarrhea that did not require hospitalization or stop the medication.

**Conclusions:** Our strategy has been associated with a reduction in the burden of hospital pressure, and it seems to be successful in terms of the number of patients who have developed serious complications and / or death. None of the patients died in the studied period and only 6 have to be admitted in conventional hospitalization area.

## INTRODUCTION

COVID-19 is an emerging zoonotic disease caused by the new bat-related coronavirus named Severe Acute Respiratory Syndrome Coronavirus 2 (SARS-CoV-2) that began at the end of 2019 in China (1). To date, SARS-CoV-2 is producing a pandemic with a high morbidity and mortality all over the world, and at the time of the submission of this paper there are at least 4,357,567 infected people in the world with 293,226 dead in the world and 269,520 infected in Spain and 26,920 dead (2).

Most of patients with SARS-CoV-2 infection are asymptomatic or present mild symptoms without clinical complications, but it is estimated that at least 20% could require hospital admission and up to 6.1% of infected patients can develop severe disease or die (3, 4). At the time of writing this paper no medical therapy is officially recommended or has shown results in improving the outcomes in COVID-19 patients (5). An easy-to-use strategy based on an oral administration regimen outside the hospital could be the one based on the use of hydroxychloroquine (HCQ) plus azithromycin (AZM) for a short period of time. HCQ with or without AZM has proven effective in small clinical trials (6, 7, 8) in different scenarios, although it has generated controversy due to the results obtained by other research groups (9).

In an attempt to decrease severe complications and the number of hospital admissions, at a time when our healthcare system was very compromised with risk of hospital collapse, we planned to implement, in the emergency room, a therapeutic strategy with at home hospital clinical monitoring service, named Hospital Home-Care Unit (HHCU), as shown in figure 1. This therapeutic strategy was offered to patients who were classified as “moderate COVID-19” (most of them with pneumonia) or were aged 55 and older. These patients were invited to follow a home care monitoring model and to be treated receiving a HCQ plus AZM based regime.

Here we present the results of this observational study (our strategy), which, although carried out in patients with mild COVID-19, seems to be safe, associated with a reduction in the burden of hospital pressure, and probably very successful in terms of the number of patients who have developed serious complications and / or death. None of the patients that followed the proposed strategy died over the period under study.

## STUDY PLANNING AND PATIENTS IDENTIFICATION

Patients ≥ 18 years old visiting the emergency room at the Hospital Universitario San Pedro de Logroño (La Rioja, Spain)* between March, 31^st^ and April, 12^th^ diagnosed with COVID-19 with confirmed SARS-CoV-2 infection by a specific PCR in a respiratory sample. Patients were classified according to the protocol shown in Figure 1, and those who met the following requirements were included in this strategy.

1. Patients with pneumonia (CURB ≤ 1) who did not present severe comorbidities and had no processes that contraindicated this therapeutic regime. To consider that a patient had pneumonia, they had to present some of the radiological findings described in COVID-19 reported by a radiologist (10).
2. Olygosimptomatic patients without pneumonia aged ≥ 55 years.
3. Patients ≥ 18 years old without pneumonia with comorbidities such as diabetes; hypertension; obesity; heart disease; nephropathy; liver disease; asthma or obstructive pulmonary disease, malignancy, HIV infection, patients in chronic treatment with immunosuppressors from any cause and pregnant women that have not met exclusion criteria.

**Figure 1:**
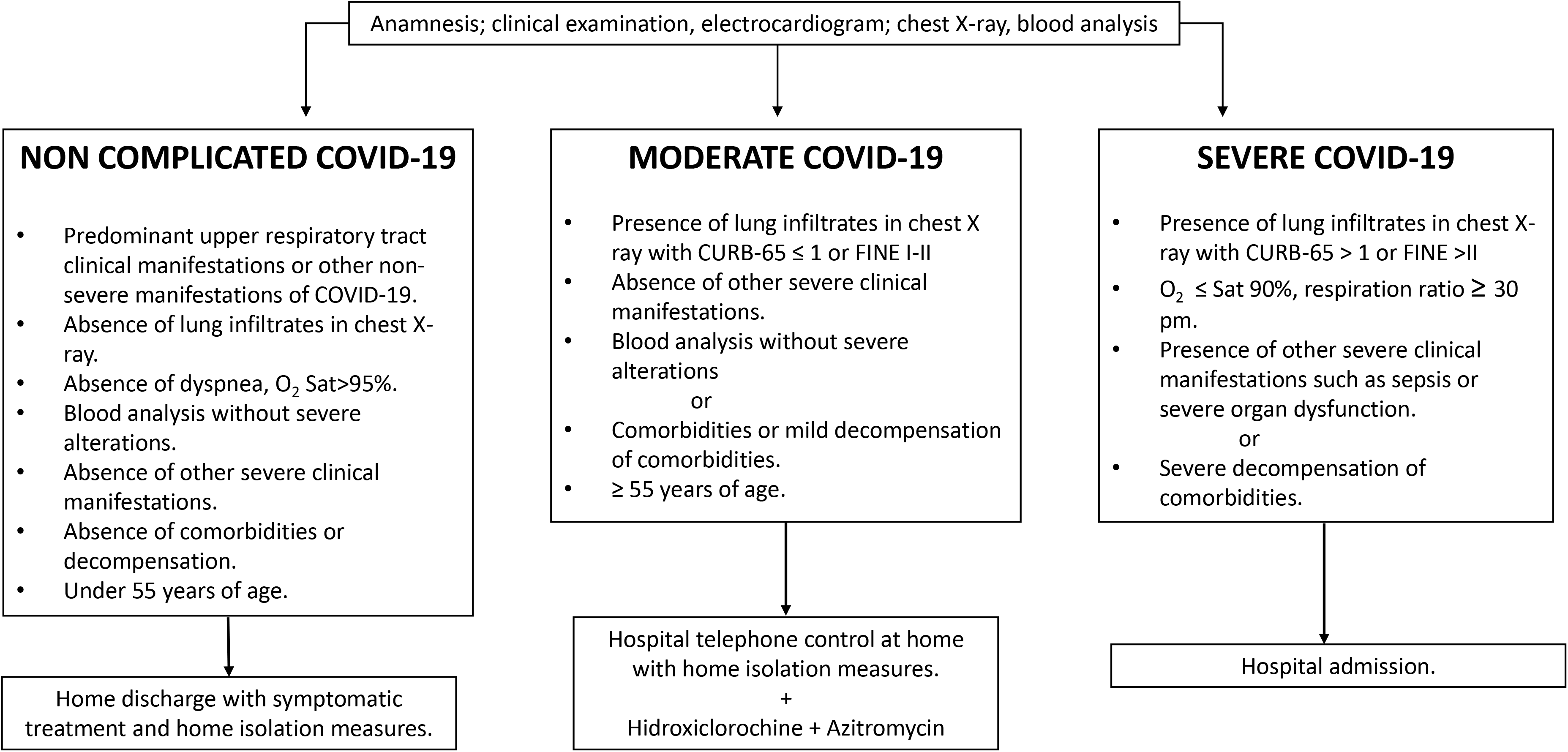
Management of COVID-19 at emergency room.

**Figure 2:**
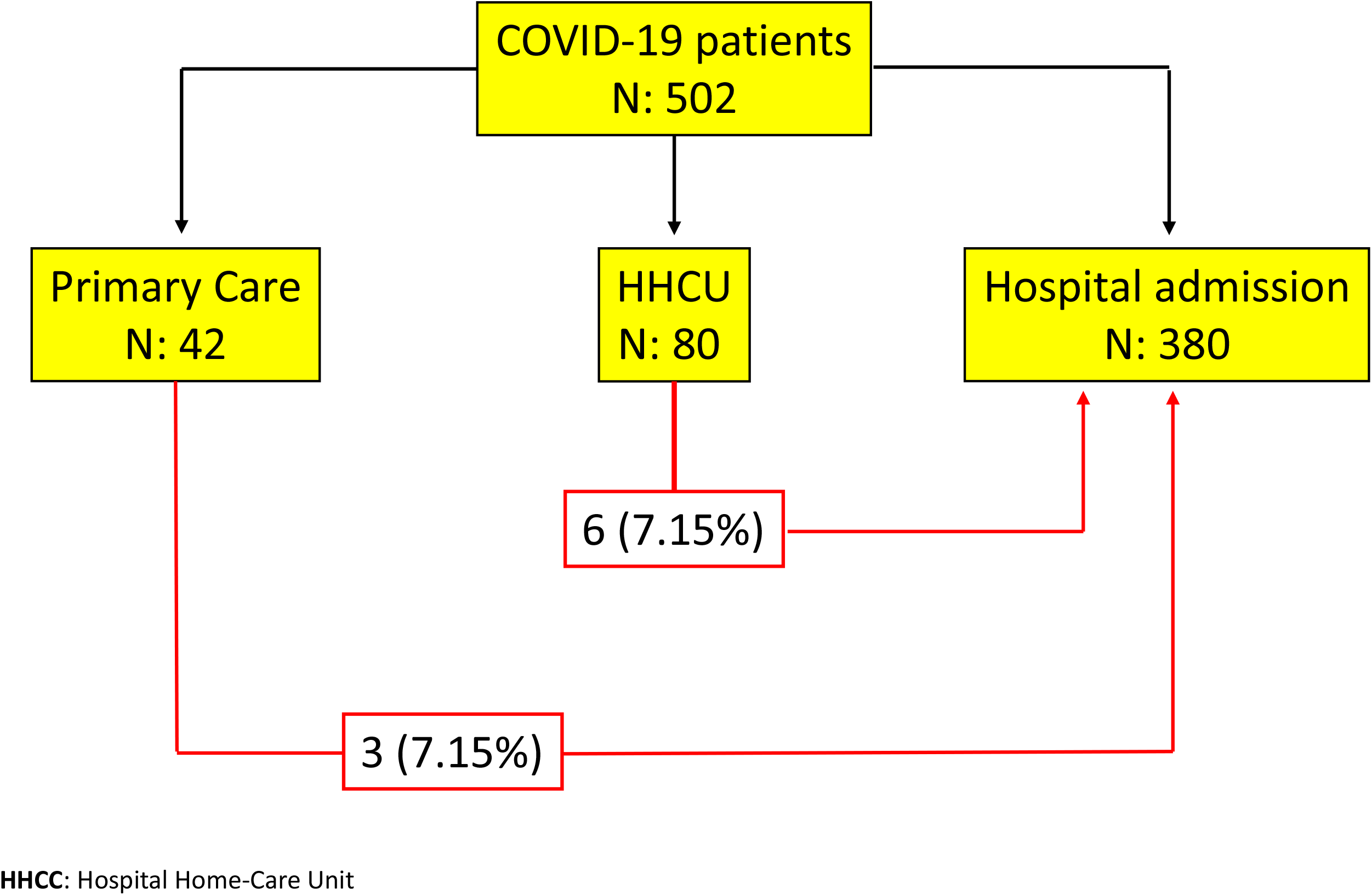
Distribution of COVID-19 patients attended in the emergency room at Hospital San Pedro in the 13 days period study.

We excluded patients with known allergies to some of the antimicrobials used. Patients treated with other drugs that increase the QTc such as antiarrhythmics or levofloxacin and those with a QTc >450msc in the electrocardiogram. Patients with retinal pathology, known G-6-PD deficiency and chronic kidney disease (phase 4 or 5 or on dialysis) were also excluded.

* Hospital Universitario San Pedro is a regional reference teaching Hospital that covers an area of 316,000 inhabitants in La Rioja (northern Spain).

## EXPOSURE (MEDICATION REGIMEN) AND DATA COLLECTION

HCQ 400 mg twice in a loading dose followed by 200 mg twice for 5 days, plus AZM 500 mg on the first day followed by 250 mg daily for 5 days.

Patients included in the strategy received a bag containing a sheet with instructions on how to take the medication and the doses of each drug according to the referred protocol and instructions about the isolation measures at home.

A daily telephone follow-up was carried out from the hospital by the same physician (involved in the fight against COVID-19) asking about adverse effects of the medication and/or clinical manifestations suggestive of disease progression, indicating in case of doubt that the patient should come again to the emergency room.

The end-points of our study were:

1. To measure the need for hospital admission within 15 days after the start of treatment.
2. To measure the need to be admitted to the intensive care unit (ICU) within 15 days after the start of the treatment.
3. To describe the severity of the clinical complications developed.
4. To measure the mortality within 30 days after starting treatment (differentiating if the cause is COVID-19 or something else).
5. To describe the safety and adverse effects of the therapeutic regime.

The data presented here, as well as other useful data for the discussion of our results, have been prospectively obtained by the physicians in charge of the patients and from the Clinical Documentation Department of the Hospital.

## RESULTS

The main characteristics of the patients attended in the emergency room at Hospital U. San Pedro during the period of the study (13 days) are shown in table 1. A total of 502 patients were attended in the emergency room due to COVID-19. Forty-two (average age 42) were sent at home under surveillance by Primary Care; 80 (average age 52) were attended by the HHCU (patients on this study) and 380 (average age 71) were admitted to the Hospital with a mortality rate of 13% of hospital admissions (non HHCU) in the studied period.

**Table 1:** characteristics of the patients attended at emergency room during the study period.

|  | Primary Care | HHCU | Hospital admission |
| --- | --- | --- | --- |
| Patients | 42 | 80 | 380 |
| Average age (limits) and median | 47 (27-76) 44 | 52 (22 to 75) 50 | 71 (26-101) 73 |
| Gender | 14 (33%) M / 28 (67%) F | 38 (47%) M /42 (53%) F | 193 (51%) M / 187 (49%) F |
| Pneumonia | 0 | 69 (85.18%) | 360 (94,73%) |
| Comorbidities | 0 | 26 (32.5%) | 241 (63.42%) |

**HHCU:** Hospital Home-Care Unit; **M:** male; **F:** female

The main characteristics of the patients are shown in table 2. From the 80 patients who were attended in HHCU, 38 (47 %) were male and 42 (53 %) female. In our series there were a group of 69 (85.18%) patients diagnosed with pneumonia (37 males and 32 females). Most of them, 57 (82.60%) had a CURB65 score of <1 (average age 49) and 12 (17.40%) a CURB score of 1 (average age 63). Eighteen (22.50%) of the pneumonia patients also had some morbidity as a risk factor: 14 of them had one comorbidity, 3 two comorbidities and 1 four comorbidities. These comorbidities were more frequent in males (61.11%) than in females (38,89%). Hypertension was present in 8 patients; diabetes mellitus in 4; cardiovascular disease in 3; chronic pulmonary disease in 3, immunosuppression or active neoplastic disease in 3; morbid obesity in 2; and liver disease in 1 patient. Other group of 11 patients (13.75%) without pneumonia were admitted to the HHCU because comorbidities or age ≥ 55 years.

**Table 2:** characteristics of the studied patients admitted to the Hospital Home-Care Unit.

| N: 80 | Total (%) | Male | Female | < 7 days from the first clinical manifestations | ≥ 7 days from the first clinical manifestations |
| --- | --- | --- | --- | --- | --- |
| Average age (limits) and median | 52 (22 to 75) 50 | 51 (30 to 75) 50 | 50 (21 to 75) 50 | 34 (42.50) | 46 (57.50) |
| ≥ 55 years old (%) | 29 (36.25) | 13 (44.82) | 16 (55.18) | 11 (37.93) | 18 (62.07) |
| Gender (%) |  | 38 (47%) | 42 (53%) |  |  |
| Pneumonia (%) | 69 (85.18) | 37 (53.62) | 32 (46.38 ) | 29 (42.02) | 40 (57.98) |
| • CURB65 < 1 (%) | 57 (82.60) | 30 (52.63) | 27 (47.37) | 23 (40.35) | 34 (59.65) |
| • CURB65 ≥ 1 (%) | 12 (17.40) | 7 (58.33) | 5 (41.67) | 6 (50%) | 6 (50%) |
| • Pneumonia + Comorbidities (%) | 18 (22.50) | 11 (61.11) | 7 (38.89) | 10 (55.55) | 8 (44.45) |
| Comorbidities or age as isolated factor without pneumonia(%) | 11 (13.75) | 1 (9.09) | 10 (90.91) | 5 (45.45) | 6 (54.55) |
| • Age as an isolated factor | 3 | 0 | 3 | 1 | 2 |
| • Hypertension | 1 | 0 | 1 | 1 | 0 |
| • Diabetes | 1 | 0 | 1 | 1 | 0 |
| • Cardiovascular disease | 0 | 0 | 0 | 0 | 0 |
| • Obesity + Chronic pulmonary disease | 1 | 1 | 0 | 1 | 0 |
| • Malignancy | 0 | 0 | 0 | 0 | 0 |
| • Chronic liver disease | 0 | 0 | 0 | 0 | 0 |
| • Chronic pulmonary disease or asthma | 3 | 0 | 3 | 0 | 3 |
| • Immunosuppressed or immunodeficiency | 2 | 0 | 2 | 1 | 1 |

Six patients with pneumonia (7.50% from the total and 8.69% of pneumonia patients group with an average age 44, 3 males and 3 females) had to be hospitalized during the observation period, 3 of them because side effects (see below) and 3 because of clinical worsening. One of these patients, a 50-year-old health-care worker woman with morbid obesity and asthma, with a 4-day clinical course from the onset of symptoms that started HCQ plus AZM the day before the admission, had clinical worsening needing mechanical ventilation at ICU because of ADRS (acute distress respiratory syndrome). With the exception of the patient admitted to the ICU, the rest of the patients were discharged at home in the following 8 days (3 to 8 days) because of clinical improvement. From 6 patients admitted to the Hospital, 5 had less than 7 days of evolution from the beginning of the clinical manifestations. This data is shown in Table 3.

**Table 3:** characteristics of patients hospitalized from the HHCU.

|  | Sex | Age | Days from the first clinical manifestations to HHCU | Clinical Diagnose | Comorbidities | ICU | Days from HHCU to hospitalization | Days from hospitalization to discharge |
| --- | --- | --- | --- | --- | --- | --- | --- | --- |
| Patient 1 | M | 51 | 4 | Admission because of QTc lengthening.<br>Pneumonia CURB 0. Diarrhea | HTA | NO | 3 | 3 |
| Patient 2 | F | 37 | 6 | Admission because of persistent cough and nausea.<br>Pneumonia CURB65 0 | WPW | NO | 9 | 4 |
| Patient 3 | F | 50 | 4 | ADRS | Morbid obesity,<br>Asthma | YES | 4 | >30 |
| Patient 4 | M | 59 | 9 | Admission because of need of O2 therapy<br>Pneumonia CURB65 0. Vomiting | NO | NO | 3 | 8 |
| Patient 5 | F | 46 | 6 | Admission because of dyspnea<br>Pneumonia CURB65 1 | Asthma | NO | 2 | 4 |
| Patient 6 | M | 51 | 3 | Admission because of distal oedema<br>Pneumonia CURB65 0. | NO | NO | 1 | 5 |

**M:** male; **F:** female; **ADRS:** acute distress respiratory syndrome. **HHCU:** Hospital Home-Care Unit; **WPW:** Wolf Parkinson White Syndrome

Twelve patients (15%), 11 of whom had pneumonia, experienced side effects which could be related to the prescribed medication. Side effects mainly affected the digestive tract with diarrhea and vomiting in 6 and 4 patients. For another patient who returned to the emergency room because of malaise, a QTc interval prolongation (452 msc) was observed in the 4^th^ day of treatment. In total 3 of these patients had to be admitted in the Hospital, 2 because of vomiting and 1 because a QTc interval lengthening. None of the patients needed to stop the HCQ or AZM and all the 80 patients finished the therapeutic strategy. From the group without pneumonia only a patient developed diarrhea that did not require hospitalization or stop the medication.

Ruling out the admissions related to the adverse effects of the medication, there were no differences in the clinical evolution of the patients who had started with clinical manifestations before/after 7 days from the beginning of the disease.

As additional data, 13 days prior to starting with the HHCU strategy, 425 patients (240 male and 185 female) were admitted to the Hospital and 60 patients were sent home for control by Primary Care with no specific treatment (ie: paracetamol). Of these last patients, 3 had to be admitted to the Hospital in the following days because of clinical worsening. Patients at that time with the criteria of HHCU were admitted to the conventional hospitalization in a COVID area.

## DISCUSSION

COVID-19 has severely hit health systems in many western countries such as Spain in a short period of time. Until we have an effective vaccine, the only useful measures to contain COVID-19 are isolation and early therapeutic intervention. At the time this strategy was launched, our emergency room was in great demand and hospital bed resources were close to collapsing. For these reasons, we planned an alternative to admission in a conventional hospitalization area. Here we analysed a first short period of 13 days. In those days we sent 80 patients to the HHCU, who relieved the hospital burden, and at the time we are writing this paper, a total of 156 COVID-19 patients have been sent to HHCU. We have not had a control group to compare to at the same time but we know that in the previous weeks, all patients with pneumonia associated to COVID-19 were admitted to hospital, so, at first, our strategy has been associated with a reduction in the hospital burden with very few complications, no deaths, and few and non-severe side effects.

None of our patients have died in the 30 days of follow-up. Only 1 patient required an ICU because ADRS and the need of mechanical ventilation and 5 other patients (total 7.5%) had to be admitted to the hospital within 15 days of starting treatment. Most of them (excepting the ICU patient) were admitted because of side effects affecting the intestinal tract or clinical worsening of their pneumonia and needed oxygen. These were discharged in the next 8 days (3 to 8 days). Although 15% of the patients reported some adverse effect, most affecting the digestive tract such as nausea and vomiting, frequently associated with the use of HCQ and probably enhanced with the use of AZM (11), these clinical manifestations are not clearly related to medication (at least in all patients), since these symptoms also frequently appear in COVID-19 (12). Our data show that the combination of HCQ plus AZM seems to be safe in selected patients with the doses we have used as suggested by other authors (11).

The availability of an easy-to-take oral therapeutic regimen for short-term with known adverse effects was an indispensable condition for carrying out this strategy. HCQ is a chloroquine analogue widely used in the treatment of rheumatic diseases, with a better safety profile and easy oral administration. HCQ was shown to have *in vitro* activity against the previous SARS-producing coronavirus in China (13) and has been shown to be more effective than chloroquine *in vitro* against SARS-CoV-2, having been recommended by the Chinese authorities to treat COVID-19 at doses of 400 mg given twice daily for 1 day, followed by 200 mg twice daily for 4 more days (14). Our choice was based on this recommendation and on the experience of the Marseille Group and other work carried out in China (6, 7). The Marseille group, in early March, studied 36 patients with COVID-19 in a clinical trial, showing that the use of the HCQ seemed safe and was associated with a reduction in medical complications. They also observed a decrease / disappearance of viral load SARS-CoV-2 and that these beneficial effects were reinforced by adding AZM (7). This study has created controversies in the same country (9). Subsequently, the Marseille group published an observational study in 80 patients and the results confirmed the value of the combination of HCQ plus AZM (15). HCQ was also analysed in a randomized clinical trial in China in which HCQ was added to conventional therapy (control arm) reducing the risk of progression, the time to clinical recovery promoting the absorption of pneumonia more quickly than controls (6). Finally, another very recent article published on-line suggests that the addition of HCQ to conventional therapy is associated with a decreased mortality in critically ill patients with COVID-19 (8).

In summary, our strategy is associated with a reduction in the burden of hospital pressure, and it seems to be safe and successful in terms of the number of patients who have developed serious complications and / or death. None of the patients died in the studied period. Because of this, we have continued throughout the time with this strategy and we present here our preliminary experience that may be of help to other Health-Care Centres, which are subject to a great deal of assistance burden. We were able to prescribe HCQ in our hospital despite the fact that HCQ was withdrawn from the market in Spain in order to avoid self-prescription and the possible adverse effects derived from misuse because of the call effect that the advertising of the clinical trials and results had on the press. In addition, this treatment regime is inexpensive, which is very interesting for developing countries.

## Data Availability

All data referred to in the manuscript are available upon request to authors. This kind of study was an authorized option in Spain, with or without a clinical trial. This is not a clinical trial. It is an observational study with an intervention approved by the Direction of the Hospital.
A few weeks ago, our healthcare system was on the verge of collapse, we were running out of hospital beds, and with the strategy of a therapeutic approach easy to carry out in a Hospitality Home-Care Unit, we were able to effectively reduce the hospital burden. We think that these data can be useful to expand knowledge in this therapeutic pattern.

## ACKNOWLEDGEMENTS

We are grateful to Dr. Aránzazu Portillo (Hospital U. San Pedro-CIBIR) for her support.

